# Prevalence of Mental Health Problems During Virus Epidemics in the General Public, Health Care Workers and Survivors: A Rapid Review of the Evidence

**DOI:** 10.1101/2020.05.19.20103788

**Authors:** Simeon J. Zürcher, Philipp Kerksieck, Christine Adamus, Christian Burr, Anja I. Lehmann, Flavia K. Huber, Dirk Richter

## Abstract

**Background:** The swift spread of SARS-CoV-2 provides a challenge worldwide. As a consequence of restrictive public health measures like isolation, quarantine, and community containment, the provision of mental health services is a major challenge. Evidence from past virus epidemics and the current SARS-CoV-2 outbreak indicate high prevalence rates of mental health problems (MHP) as short- and long-term consequences. However, a broader picture of MHP among different populations is still lacking.

**Methods:** We conducted a rapid review on MHP prevalence rates published since 2000, during and after epidemics, including the general public, health care workers, and survivors. Any quantitative articles reporting on MHP rates were included. Out of 2855 articles screened, a total of 74 were included in this review.

**Results:** Most original studies on MHP were conducted in China in the context of SARS-CoV-1, and reported on anxiety, depression, post-traumatic stress symptoms/disorder, general psychiatric morbidity, and psychological symptoms. The MHP rates across studies, populations, and epidemics vary substantially. While some studies show high and persistent rates of MHP in populations directly affected by isolation, quarantine, threat of infection, infection, or life-threatening symptoms (e.g. health care workers), other studies report minor effects. Furthermore, even less affected populations (e.g. distant to epidemic epicenter, no contact history with suspected or confirmed cases) can show high rates of MHP.

**Discussion:** MHP vary largely across countries and risk-groups in reviewed studies. The results call attention to potentially high MHP during epidemics. Individuals affected directly by an epidemic might be at a higher risk of short or even long-term mental health impairments. This study delivers insights stemming from a wide range of psychiatric instruments and questionnaires. The results call for the use of validated and standardized instruments, reference norms, and pre-post measurements to better understand the magnitude of the MHP during and after the epidemics. Nevertheless, emerging MHP should be considered during epidemics including the provision of access to mental health care to mitigate potential mental impairments.

## 1 Introduction

In the past two decades, many countries faced challenges in the realm of major infectious disease epidemics including SARS-CoV-1 (Peiris et al., 2003), Swine flu (H1N1) (Trifonov et al., 2009), Middle East respiratory syndrome coronavirus (MERS-CoV) (Zaki et al., 2012), avian influenza (H7N9) (Gao et al., 2013), Ebolavirus (Baseler et al., 2017), and the recent worldwide SARS-CoV-2 outbreak (Wu et al., 2020). Epidemic outbreaks can result in high case fatality rates and morbidity (Van Bortel et al., 2016; Meo et al., 2020) and may require communities to introduce restrictive public health measures like isolation, mass quarantine, and community containment interventions in order to stop transmissions and save lives (Wilder-Smith and Freedman, 2020). In consequence, epidemics can cause a high individual and societal burden and can lead to substantial economic loss (Smith, 2006; Mak et al., 2009; Van Bortel et al., 2016; Dorn et al., 2020). While considerable efforts rely on protective and treatment measures such as virus transmission pathways, clinical presentations, and the development of vaccinations, attention is only recently given to short or long-term mental health problems (MHP, hereafter defined as psychiatric/psychological symptoms and mental illness/disorders) (Rajkumar, 2020) that may arise due to the different surrounding consequences of an epidemic in the general public, health care workers (HCW), and survivors of infectious diseases (survivors).

Epidemics can negatively impact a substantial part of the general public in many different ways such as feelings of a personal threat of being infected (Van Bortel et al., 2016; Brooks et al., 2020; Chew et al., 2020), worries about relatives and family members or losing loved ones (Brooks et al., 2020; Chew et al., 2020; Li et al., 2020), and protective measures like mass quarantining, the consequences of which leads to individual and social restrictions, and economic loss (Brooks et al., 2020). As a result, these factors can elicit feelings of anxiety, anger, loneliness, grief, boredom and may lead to serious MHP (Brooks et al., 2020; Chew et al., 2020; Fardin, 2020). Furthermore, the extensive and sometimes controversial mass media coverage during epidemics may amplify uncertainty, loss of control and anxiety (Brooks et al., 2020; Fardin, 2020). Aside from the general public, HCW are prone to different MHP since they usually face an immediate threat of infection through patient contact by working at the epidemic frontline. Studies suggest that HCW accounted for up to 57% of SARS-CoV-1, 27% of MERS-CoV, and 12% of Ebola cases in some countries, which frequently resulted in morbidity or even death (Chan-Yeung, 2004; Suwantarat and Apisarnthanarak, 2015). In HCW, epidemics often result in difficult working conditions like staff shortage, increased workload (Van Bortel et al., 2016), overwhelming patient numbers (Suwantarat and Apisarnthanarak, 2015; Van Bortel et al., 2016), limited safety equipment (Van Bortel et al., 2016), and quarantine or isolation after infectious disease transmission (Van Bortel et al., 2016; Brooks et al., 2020). Furthermore, HCW often suffer social consequences like stigma (Bai et al., 2004; Verma et al., 2004; Van Bortel et al., 2016), mistrust and violence (Van Bortel et al., 2016) avoidance from relatives, and the fear of infecting others (Verma et al., 2004). Given the high risk of transmission, HCW often account for a substantial fraction of survivors, who frequently experience isolation, intensive treatment, stigmatization and exposure to an immediate threat of morbidity or death (Van Bortel et al., 2016; Troyer et al., 2020). To date, many studies exist that describe MHP related to epidemics across a wide range of populations. However, to the best of our knowledge no review covering MHP during epidemics currently exists.

### 1.1 Objectives

The purpose of this rapid review is to provide an overview of MHP prevalence rates during and after large epidemics of the past two decades. We aim to provide a broad picture of MHP that may arise across a wide range of populations including a) the general public, b) HCW, and c) and virus disease survivors.

## 2 Materials and Methods

### 2.1 Search Strategy

The rapid and dynamic development of the current situation with SARS-CoV-2 requires quick evidence synthesis in order to inform decision-making processes in health care systems. The methodology of this article is based on the practical guide for rapid reviews provided by WHO (Tricco et al., 2017). We undertook a review of evidence on prevalence rates during and after epidemic outbreaks on MHP in the general public, HCW, and survivors. The focus was on SARS-CoV-1, H1N1, MERS-CoV, H7N9, Ebolavirus, and SARS-CoV-2. PubMed was searched on April 1, 2020 with a broad search strategy (see **Supplementary Table 1**).

### 2.2 Participants, Interventions, and Comparators

Any type of quantitative study that provided prevalence rates of MHP in adults (≥18 years) during and after epidemic outbreaks, published in English from the year 2000 to March 31, 2020 was included. Studies that measured MHP rates assessed by psychometrically validated instruments, diagnostic interview, and medical records (chart review), were also included. We excluded studies that used a qualitative design, that did not report on MHP prevalence rates (e.g. providing mean scores only), that did not provide prevalence rates based on previously defined cut-off values for a measurement instrument (e.g. median based sample splitting), and that included MHP measured by single questions/items. Studies on common seasonal influenza were also excluded. Furthermore, general states like social functioning, quality of life, generic fears (e.g. fear of contracting a virus or worries) or stigma were excluded. Based on the titles and abstracts of studies, potential eligible studies of the database search were selected by CA using a co-developed standardized review form to assess study eligibility. The primary reviewer SJZ then selected eligible studies after searching the full-text of each potentially eligible publication. Doubts and uncertainty in eligibility of a certain study were solved by discussion.

### 2.3 Data Sources, Study Selection, and Data Extraction

An electronic data extraction form was developed to assess the characteristics of the included studies and the reported MHP prevalence rates. Data was extracted by SJZ, CA, PK, and FH. Collected items included: author(s), year of publication, country or region, number of participants, type of epidemic outbreak, time point of assessment, type of MHP assessed, MHP prevalence rate, and assessment method. Time point of assessment was coded as: during epidemic/hospital stay, post-epidemic/discharge including one-year follow-up (≤1y), between one and four years follow-up (>1-4y), or a combination of both if applicable (e.g. for longitudinal studies). MHP were categorized into anxiety, depression, post-traumatic symptoms/disorders (PTSD) or stress, burnout, psychiatric morbidity, and further MHP like hallucinations or insomnia. We used baseline assessment data for intervention studies that provided prevalence rates. Data was stratified by the following populations: a) general public including general surveys, b) HCW including all hospital staff, military duty members, and family members as caregivers involved in active treatment or in potential contact with patients, and c) infectious disease survivors (that may include suspected cases in some studies). Data quality and strength of evidence was not rated in the current review. All authors who extracted data discussed possible uncertainties with the primary reviewer SJZ.

### 2.4 Data Analysis

Included studies varied in assessment of MHP (e.g. questionnaires, diagnostic interviews), MHP instruments with applied cut-off scores, sampling methods and response rates, outbreak-related time points of assessments, and in regional differences in the magnitude/level of affect. Due to the approach chosen (rapid review), no meta-analysis was conducted. Therefore, a descriptive approach was utilized to synthesize reported MPH prevalence rates. If provided, we show MHP rates from a moderate degree of severity as defined by authors within original studies.

## 3 Results

### 3.1 Study Characteristics

Our PubMed search yielded 2,855 articles of which 74 were included in the qualitative synthesis (see **Figure 1**). The majority of studies were cross-sectional in design and focused on MHP during SARS-CoV-1 (n = 41), followed by Ebolavirus (n = 12), MERS-CoV and SARS-CoV-2 (n = 7), H1N1 (n = 6), and H7N9 (n = 1). About half of the studies in the general public used random sampling, while the majority of articles in HCW and survivors were non-random samples. The vast majority of studies was conducted in China, including Taiwan and Hong Kong (n = 39), followed by other countries in Asia (n = 14), in Africa (n = 12), and the American continent (mainly Canada; n = 6), with three studies conducted in Europe. We found n = 28, 26, and 20 studies that investigated the general public, HCW, and s0urvivors, respectively. The vast majority of studies assessed MHP using self-reported questionnaires, while only few used standardized diagnostic interviews. Results stratified by general public, HCW, and survivors can be found in **Tables 1–3**.

**Figure 1.**
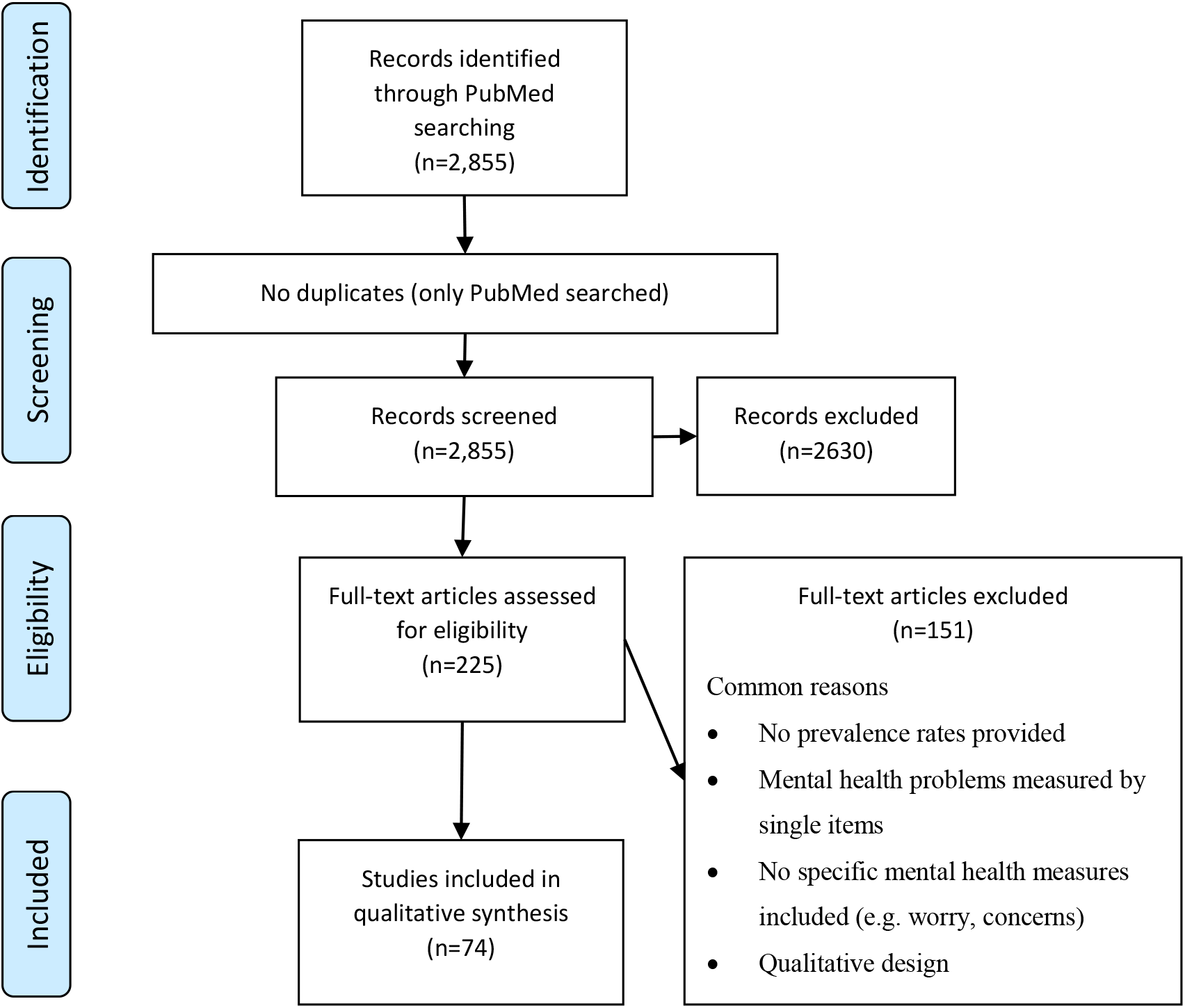
PRISMA flow diagram of the studies retrieved for the review

**Table 1.** Prevalence rates of mental health problems in the general public in chronological order of the respective epidemic outbreak

| Author (year) | Country /Region | N (including other population) | Epidemic / Time point | Mental health problem<sup>a</sup> (Assessment instrument<sup>b</sup>): Prevalence rates |
| --- | --- | --- | --- | --- |
| Cao et al. (2020) | China | 7143 | SARS-CoV-2 / During | **Anxiety** (GAD-7): 2.7% moderate; 0.9% severe |
| Qiu et al. (2020) | China, Hong Kong, Taiwan | 52730 | SARS-CoV-2 / During | **PTSD/Stress** (CPDI): 29.9% mild to moderate; 5.14% severe |
| Wang et al. (2020a) | China | 1210 | SARS-CoV-2 / During | **Anxiety** (DASS-21): 20.4% moderate; 8.4% severe/extremely severe<br>**Depression** (DASS-21): 12.2% moderate; 4.3% severe/extremely severe<br>**PTSD/Stress** (IES-R): 53.8% moderate/severe<br>**PTSD/Stress** (DASS-21): 5.5% moderate; 2.6% severe/extremely severe |
| Wang et al. (2020b) | China | 600 | SARS-CoV-2 / During | **Anxiety** (SAS): 0.67% moderate; 0% severe<br>**Depression** (SDS): 2.5% moderate; 0.33% severe |
| Zhang and Ma (2020) | China | 263 | SARS-CoV-2 / During | **PTSD/Stress** (IES): 7.6% moderate/severe |
| Kamara et al. (2017) | Sierra Leone | 143 | Ebolavirus / During | **Further MHP** (Med-rec): <1% alcohol/substance use disorder; 21% psychotic symptoms (e.g. hallucinations); 12% moderate to severe emotional disorder or depression |
| Koroma et al. (2019) | Sierra Leone | 10011 | Ebolavirus / During / Post ≤1 y | **Further MHP** (Med-rec), any mental health disorders in various hospital types: <1% pre-Ebola; <1% during Ebola; ≤1% post-Ebola |
| Betancourt et al. (2016) | Sierra Leone | 1008 | Ebolavirus / Post ≤1 y | **Anxiety** (HSCL-25): 1.3%<br>**Depression** (HSCL-25): 1.4%<br>**PTSD/Stress** (PSS-I): 11.3% likely PTSD |
| Jalloh et al. (2018) | Sierra Leone | 3564 | Ebolavirus / Post ≤1 y | **Anxiety/Depression** (PHQ-4): 48.6% any symptoms<br>**PTSD/Stress** (IES-R): 76.4% any symptoms; 27% levels of clinical concern for PTSD; 16% levels of probable PTSD diagnosis |
| Mollers et al. (2015) | Netherlands | 72 | MERS-CoV / During | **PTSD/Stress** (IES-R): 22% |
| Al-Rabiaah et al. (2020) | Saudi Arabia | 174 | MERS-CoV / During | **Anxiety** (GAD-7): 4.6% moderate; 0% severe |
| Jeong et al. (2016) | Republic of Korea | 1'692 (incl. HCW, Survivors) | MERS-CoV / Post ≤1 y | **During isolation:**<br>**Anxiety** (GAD-7): 47.2% MERS positive; 7.6% negative<br>**Further MHP/Anger** (STAXI): 52.8% MERS positive; 16.6% negative<br>**4-6 months after isolation:**<br>**Anxiety** (GAD-7): 19.4% MERS positive; 3% negative<br>**Further MHP/Anger** (STAXI): 30.6% MERS positive; 6.4% negative |
| Rubin et al. (2009) | England, Scotland | 997 | H1N1 / During | **Anxiety** (STAI-6): 23.8% symptoms; 2.1% high |

**Table 1** Continued
|  |  |  |  |  |
| --- | --- | --- | --- | --- |
| Wang et al. (2011) | China | 419 | H1N1 / During | **Psychiatric morbidity** (SRQ-20): 8% quarantined group; 14% non-quarantined group<br>**PTSD/Stress** (IES-R): 10.8% quarantined group; 16.9% non-quarantined group |
| Xu et al. (2011) | China | 1082 | H1N1 / During | **PTSD/Stress** (PCL-C): 2% symptomatic PTSD |
| Leung et al. (2003) | Hong Kong | 1115 | SARS-CoV-1 / During | **Anxiety** (STAI): 12.6% quite/very anxious |
| Hawryluck et al. (2004) | Canada | 129<br>(incl. HCW) | SARS-CoV-1 / During | **Depression** (CES-D): 31.2%<br>**PTSD/Stress** (IES-R): 28.9% |
| Quah and Hin-Peng (2004) | Singapore | 1202 | SARS-CoV-1 / During | **Anxiety** (CAS): 42.4% moderate; 2.9% high |
| Lau et al. (2005) | Hong Kong | 818 | SARS-CoV-1 / During | **PTSD/Stress** (IES): 13.3% males, 18.0% females moderate to severe; 1.3% males, 1.5% females severe |
| Lau et al. (2006) | Hong Kong | 818 | SARS-CoV-1 / During | **PTSD/Stress** (IES): 16% moderate to severe |
| Lee et al. (2006a) | Hong Kong | 235 | SARS-CoV-1 / During | **Depression** (BDI): 12.3% |
| Chan et al. (2007) | Hong Kong | 122 | SARS-CoV-1 / During | **Anxiety** (STAI): 29.5% moderate; 4.1% high |
| Reynolds et al. (2008) | Canada | 1057<br>(incl. HCW) | SARS-CoV-1 / During | **PTSD/Stress** (IES-R): 14.6% |
| Sim et al. (2010) | Singapore | 415 | SARS-CoV-1 / During | **Psychiatric morbidity** (GHQ-28): 22.9%<br>**PTSD/Stress** (IES-R): 25.8% high levels |
| Ko et al. (2006) | Taiwan | 1499 | SARS-CoV-1 / Post ≤1y | **Depression** (TDQ): 3.7% depressive symptoms |
| Lee et al. (2006b) | Hong Kong | 146 | SARS-CoV-1 / Post ≤1y | **Depression** (CES-D): 32.4% elderly; 18.7% middle-aged<br>**PTSD/Stress** (IES-R): 14.1% elderly; 4% middle-aged |
| Mihashi et al. (2009) | China | 187 | SARS-CoV-1 / Post ≤1y | **Psychiatric morbidity** (GHQ-30): 24.6% during the isolation period; 26.2% during the recovery period |
| Peng et al. (2010) | Taiwan | 1278 | SARS-CoV-1 / Post ≤1y | **PTSD/Stress** (BSRS-5): 11.7% |

<sup>a</sup> PTSD, Post-traumatic stress disorder
<sup>b</sup> BDI, Beck Depression Inventory; BSRS-5, 5-item Brief Symptom Rating Scale; CAS, B.A. Thyer's Clinical Anxiety Scale; CES-D, Center for Epidemiological Studies Depression Scale; CPDI, COVID-19 Peritraumatic Distress Index; DASS-21, Depression, Anxiety and Stress Scale; GAD-7, 7-item Generalized Anxiety Disorder Scale; GHQ, General Health Questionnaire; HSCL-25, Hopkins Symptom Checklist-25; IES-R: Impact of Event Scale – Revised; Med-rec, medical records; PCL-C, PTSD Checklist – Civilian Version; PHQ-4, Patient Health Questionnaire; PSS-I, PTSD Symptom Scale-Interview; SAS, Self-Rating Anxiety Scale; SDS, Self-Rating Depression Scale; SRQ-20, Self-Report Questionnaire; STAI, Spielberger State-Trait Anxiety Inventory; STAXI, State-Trait Anger Expression Inventory; TDQ, Taiwanese Depression Questionnaire

**Table 2.**
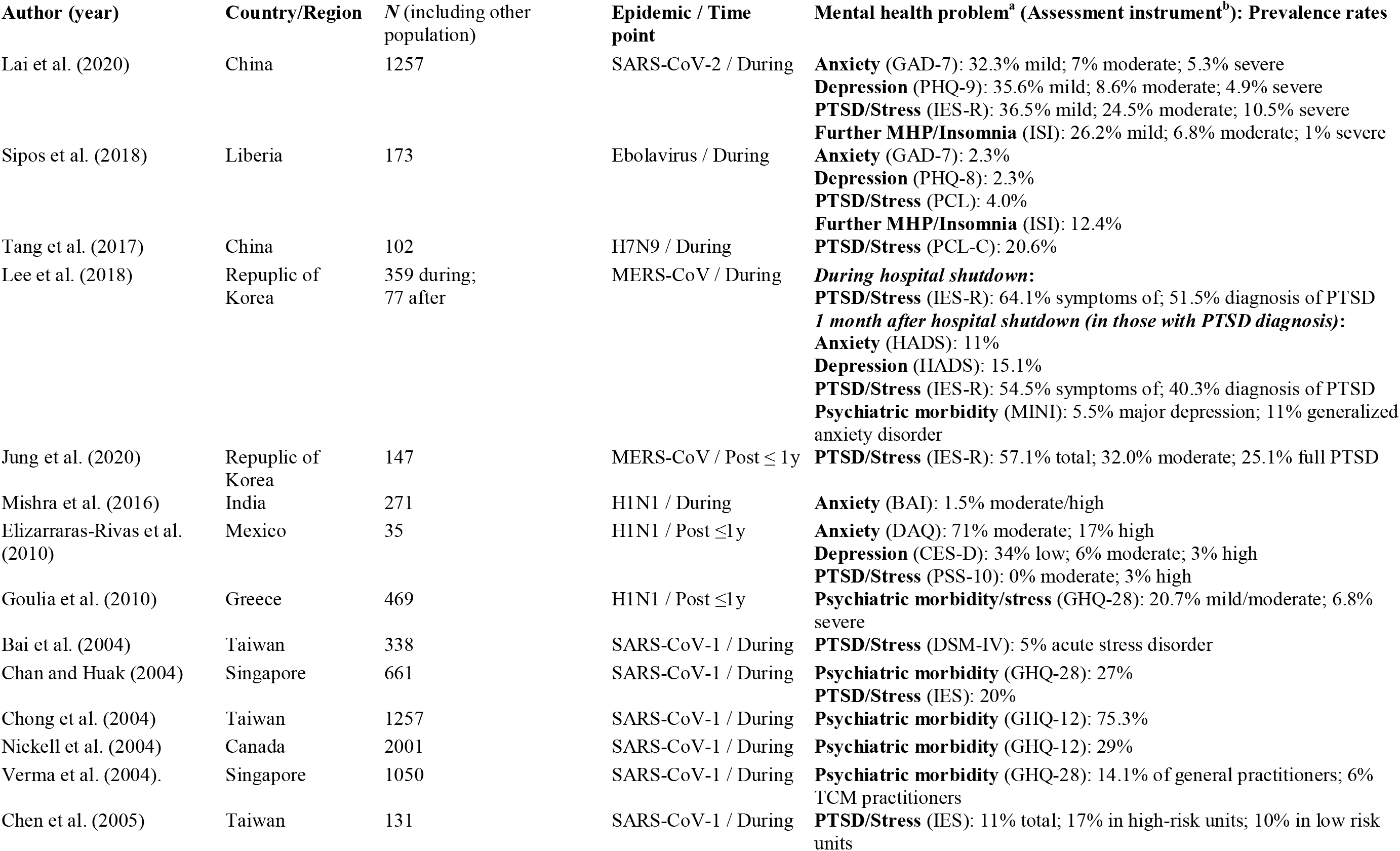
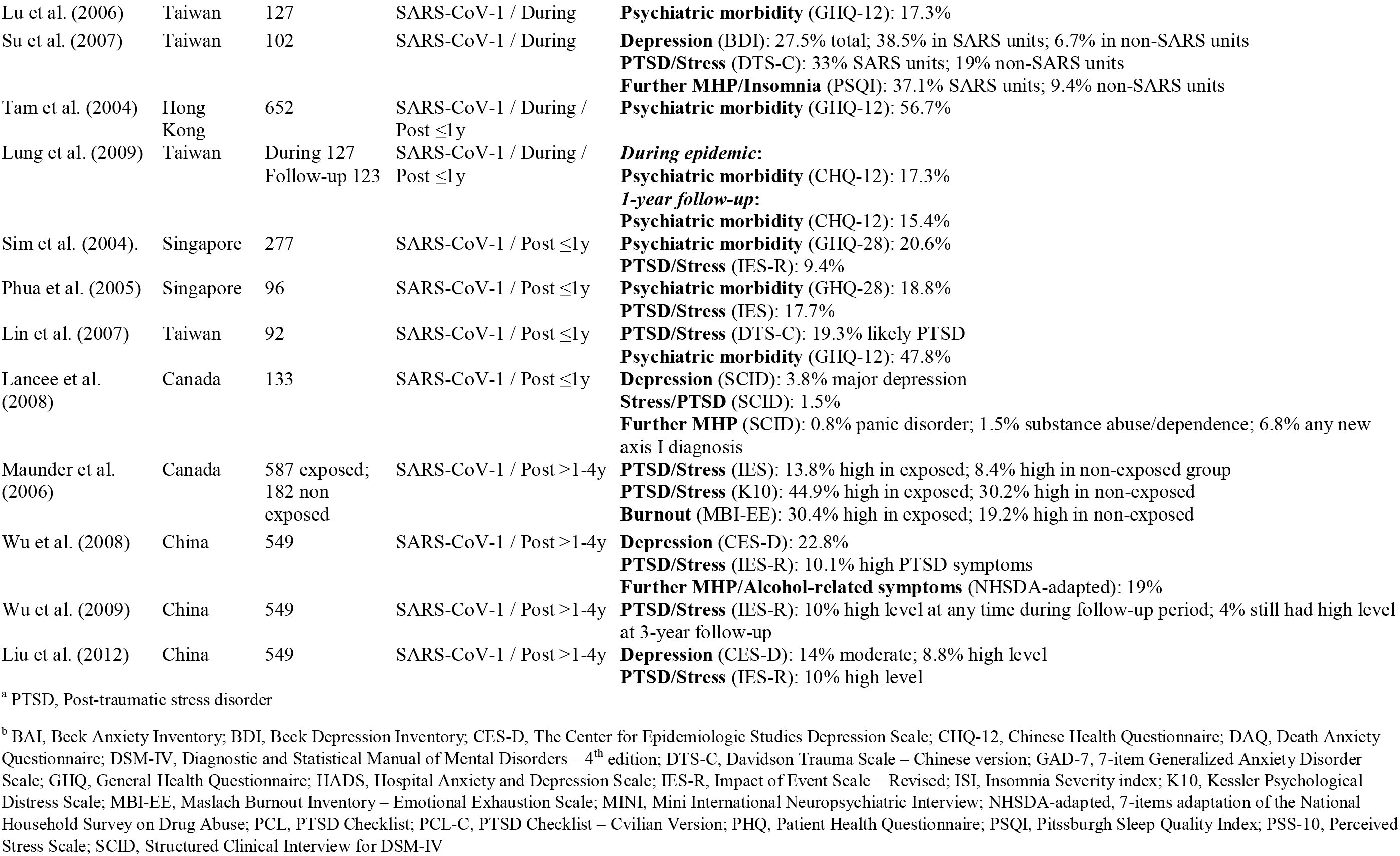
Prevalence rates of mental health problems in health care workers in chronological order of the respective epidemic outbreak

**Table 3.**
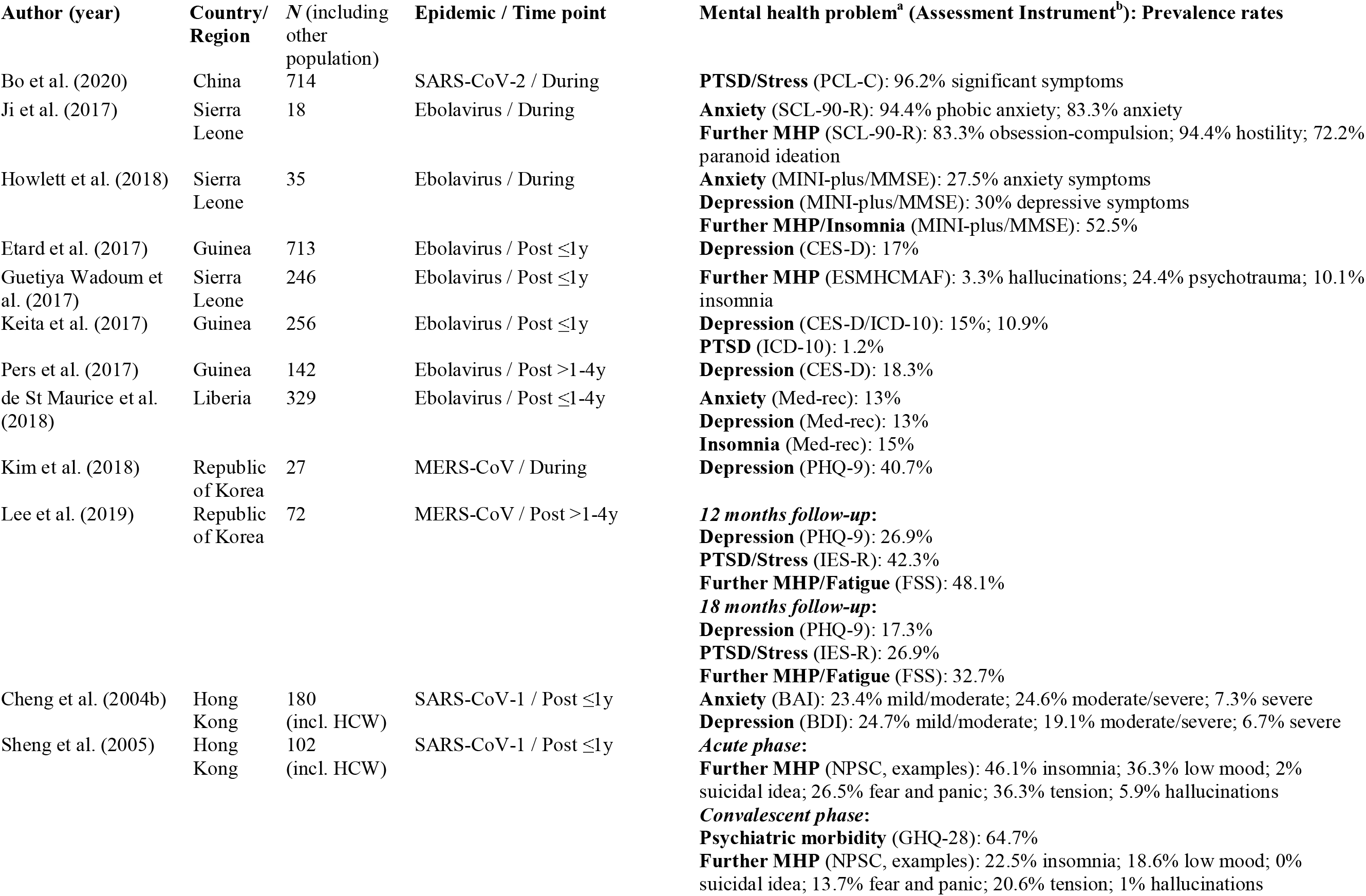
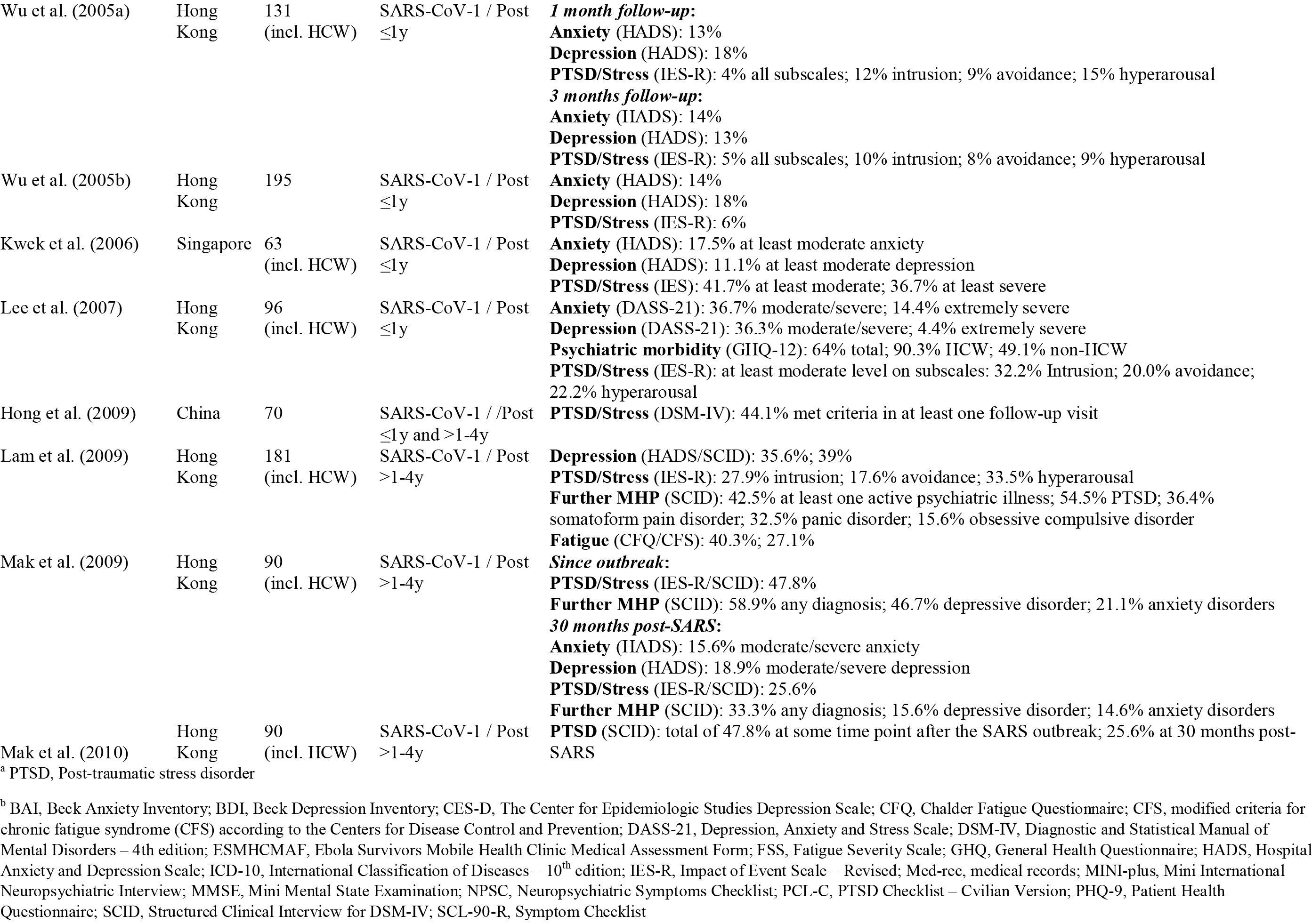
Prevalence rates of mental health problems in health care workers in chronological order of the respective epidemic outbreak

### 3.2 Synthesized Findings

#### 3.2.1 General public

Range of prevalence rates across original articles were as follows: anxiety (0.7–47.2%), depression (1.4–32.4%), any anxiety/depression symptoms combined (48.6%), PTSD/stress (2.0–76.4%), and psychiatric morbidity (8.0–26.2%). The rates of further MHP included any mental disorder (<1.0%), alcohol/substance use disorders (<1.0%), anger (6.4–52.8%), moderate to severe emotional disorder or depression (12.0%), intellectual disability (5.0%), and psychotic symptoms like hallucinations (21.0%). The highest and lowest rates of anxiety were found in MERS-CoV (48.6%), and SARS-CoV-2 (0.7%), respectively. For depression the highest rates were found in SARS-CoV-1 (32.4%) and the lowest in Ebolavirus (1.4%). For PTSD/stress, the highest rates were shown for Ebolavirus (76.4%), and the lowest in H1N1 (2.0%). Psychiatric morbidity was highest in SARS-CoV-1 (26.2%) and lowest in H1N1 (8.0%).

#### 3.2.2 Health care workers

Range of prevalence rates were as follows: anxiety (1.5–88.0%), depression (2.3–49.1%), PTSD/stress (1.5–71.5%), burnout (19.2–30.4%), and psychiatric morbidity (6.0–75.3%). The rates of further MPH included any new Axis 1 diagnosis (6.8%), insomnia (9.4–37.1%), and substance abuse or alcohol related symptoms (1.5%–19.0%). The full range of rates in anxiety were both found in H1N1 (1.5–88.0%). For depression, the highest rates were found in SARS-CoV-2 (49.1%) and the lowest in Ebolavirus (2.3%). For PTSD/stress, the highest rates were shown for SARS-CoV-2 (71.5%) and the lowest for SARS-CoV-1 (1.5%). Highest and lowest rates for psychiatric morbidity were both found for SARS-CoV-1 (6.0–75.3%).

#### 3.2.3 Survivors

Range of prevalence rates were as follows: anxiety (13.0–94.4%), depression (11.0–50.5%), PTSD/stress (1.2–96.2%), and psychiatric morbidity (49.1–90.3%). Furthermore, the rates of further MHP included any psychiatric diagnosis (33.3–58.9%), fatigue (27.1–48.1%), fear and panic (13.7–26.5%), hallucinations (1–5.9%), insomnia (10.1–52.5%), low mood (18.6–36.3%). obsession-compulsion (15.6–83.3%), panic disorder (32.5%), paranoid ideation (72.2%), somatoform pain disorder (36.4%), suicidal ideation (2.0%), and tensions/hostility (20.6–94.4%). The highest and lowest rates of anxiety were fund in Ebolavirus (94.4%), and SARS-CoV-1/Ebolavirus (13%) respectively. Depression was highest in SARS-CoV-1 (50.5%) and lowest in ebolavirus (11%). For PTSD/stress, the highest rates were shown for SARS-CoV-2 (96.2%) and lowest for Ebolavirus (1.2%). Psychiatric morbidity was described only in SARS-CoV-1 (49.1–90.3%).

## 4 Discussion

### 4.1 Summary of Main Findings

In this rapid review of 74 original articles we found a wide range of MHP including anxiety, depression, PTSD and stress related symptoms or disorders, psychiatric morbidity, and many further MHP like paranoid ideation, hallucinations, and insomnia that may occur in the general public, HCW or survivors during and after epidemic outbreaks. Aside from methodological issues and the large heterogeneity of original studies (e.g. poor validation, different cut-offs for case definition), MHP may be prevalent in all three populations. These problems may be substantial and can persist over time in HCW and survivors more directly affected by the epidemic threat. However, it should be noted that epidemic circumstances can also yield positive impacts on mental health like spending more time on physical activity and taking more care of one’s mental health (Lau et al., 2006).

### 4.2 General public

MHP ranged widely both across the general public and in all epidemics, which makes it difficult to estimate the magnitude and associated characteristics that may aggravate MHP. However, many studies investigated risk and protective factors of MHP. Although some controversy exists among studies, a higher level of epidemic exposure (e.g. living proximity to epidemic epicenter, contact history to high prevalent virus regions) (Lee et al., 2006b; Sun et al., 2020), hospitalization during epidemic (Sim et al., 2010), being quarantined (Ko et al., 2006), or having infected family members (Lee et al., 2006a; Xu et al., 2011; Cao et al., 2020) may aggravate MHP. Further risk factors include being female (Lee et al., 2007; Wang et al., 2011; Xu et al., 2011; Sun et al., 2020), chronic physical illness (Cheng et al., 2004b), poor self-rated health (Wang et al., 2020a), and dissatisfaction with measures controlling the virus (Wang et al., 2011). Furthermore, many studies reported problems like loneliness, boredom, anger, worries about family members (Wang et al., 2020a), and financial problems or economic loss (Chua et al., 2004; Lau et al., 2005; Mishra et al., 2016; Cao et al., 2020) that negatively interfere with mental health. In contrast, accurate health information (e.g., treatment, local outbreak situation) (Wang et al., 2020a), particular precautionary measures (e.g., hand hygiene, wearing a mask) (Wang et al., 2020a), social support (Ko et al., 2006; Lau et al., 2006; Cao et al., 2020), and appraisals and coping strategies (Cheng et al., 2004b; Chew et al., 2020) may be protective.

### 4.3 Health care workers and survivors

Similarly, HCW and survivors showed a wide range of mental health impacts. However, MHP rates in these populations may be more substantial than in the general public. HCW that were directly involved in patient care (Verma et al., 2004), working in high risk units and with infected patients (Chen et al., 2005; Maunder et al., 2006; McAlonan et al., 2007; Su et al., 2007) conscripted workers (Chen et al., 2005), or that underwent quarantine during outbreak (Bai et al., 2004; Liu et al., 2012) were found to be associated with a higher risk of MHP. Furthermore, younger age (Verma et al., 2004; Su et al., 2007), being single (Chan and Huak, 2004; Liu et al., 2012), fear of adversely affecting relatives (Maunder et al., 2003; McAlonan et al., 2007), pre-exposure to traumatic events or history of MHP (Su et al., 2007; Lancee et al., 2008; Liu et al., 2012) were also found to be associated with a higher risk of MHP. In contrast, adequate professional education and training (Maunder et al., 2006; Lancee et al., 2008; Tang et al., 2017), support from colleagues (Chan and Huak, 2004), appropriate information and communication (directives, precautionary measures, disease information) (Chan and Huak, 2004), and altruistic risk acceptance (Liu et al., 2012) were found to be protective. In survivors MHP may be aggravated by a history of mental illness (Jeong et al., 2016), the fear of permanent damage or death (Cheng et al., 2004b; Wu et al., 2005b), longer duration of quarantine (Hawryluck et al., 2004), having physical late sequelae (Pers et al., 2017), and impairment of ability to work (Lam et al., 2009). Furthermore, survivors that are HCW were shown to be more susceptible to long term MHP compared to non-HCW survivors (Cheng et al., 2004a; Lee et al., 2007).

### 4.4 Mental health problems and methodological issues

The methodological characteristics and quality of studies in assessing MHP ranges widely. We found only few studies that did not utilize a cross-sectional design without repetition. Further, most cross-sectional studies did not report any comparative data from which the change of prevalence rates due to the epidemic could be estimated. Sampling characteristics were also varying. Only about half of the studies in the general public were based on representative samples. As many studies were conducted during or shortly after the peak phase of the epidemic, results have to be regarded as acute stress reactions that do not allow for inference of longer-lasting MHP. While some authors used well established and widely used instruments and standardized diagnostic interviews (e.g. Ji et al. (2017) or Lancee et al. (2008)), others used instruments with unclear quality (e.g. Guetiya Wadoum et al. (2017)). Besides the possibility of biased results, this approach makes it challenging to identify clinically relevant cases. With respect to the application of diagnostic instruments, cut-off values might vary between countries and cultures. Therefore, a lack of validated, country-specific, cut-off values of the measurement instruments might be problematic (Jalloh et al., 2018).

### 4.5 Future Directives and Implications for Research, Policy, and Practice

#### 4.5.1 Monitoring MHP as a tool for mental health care provision

As shown by this review, MHP may be prevalent across a broad range of populations. In this vein, clinical monitoring of risk groups that are vulnerable to psychological impairments due to the current SARS-CoV-2 epidemic is essential (Pfefferbaum and North, 2020). Pfefferbaum and North (2020) pointed out, that the monitoring of psychosocial needs should assess SARS-CoV-2–related stressors, secondary adversities, psychosocial effects, and indicators of vulnerability. Besides others, routine outcome monitoring (Carlier et al., 2012) as a measurement feedback system, apps for (self-)monitoring of mood, sleep-quality, or medication adherence (Rubanovich et al., 2017), and artificial intelligence predicting relevant psychiatric outcomes (Lovejoy et al., 2019), are available for public mental health monitoring. In the best case, mental health service providers should be aided by e-monitoring during epidemics. As mentioned above, in research MHP should be assessed by standardized diagnostic interviews or measurement instruments enabling appropriate case detection identifying risk groups in order to inform policy and practice.

#### 4.5.2 Access to mental health service in epidemics

Furthermore, access to mental health services for those in need is paramount during the SARS-CoV-2 crisis, especially when social isolation is experienced (Wang et al., 2017). Beside the psychosocial consequences of public health measures such as quarantine (Brooks et al., 2020), acute viral infection is unknown but likely to be accompanied by substantial neuropsychiatric symptoms (anxiety, depression, and trauma-related symptoms) as a host immunologic response to the infection (Troyer et al., 2020). Mental health care interventions are expected to reduce symptoms such as PTSD (Torales et al., 2020). However, during epidemic scenarios care needs to be adapted to upcoming circumstances by respective governments in order to prevent or support individuals with MHP (Duan and Zhu, 2020). In epidemic conditions, where consultation in-person is restricted there are important implications for digital health approaches. Online psychotherapy and consultation might help to improve access to mental health care, particularly in times of quarantine and isolation (Langarizadeh et al., 2017; Tuerk et al., 2018). It does need to be highlighted that the effectiveness of online services for the improvement of mental health services requires further assessment (Kauer et al., 2014). Consequently, the outbreak of SARSCoV-2 calls for rapid reports and insights, as well as long-term health service research focusing on both remote and in-person mental health resources during epidemics (Starace and Ferrara, 2020; Wind et al., 2020).

#### 4.5.3 Implications for HCW as a highly demanded group

Working conditions play an important role in mental health. For HCW, protective working conditions such as social support, constructive communication and staff training and education have already been mentioned in some studies (Maunder et al., 2006; Lancee et al., 2008; Tang et al., 2017). Employers should consider strengthening these resources by implementing support systems and coping management strategies. Besides such protective factors there might be even health promoting occupational aspects to be considered. For HCW, the intent to help can buffer mental health-impairing consequences (Liu et al., 2012) but might be a rewarding factor in and of itself (De Gieter et al., 2006). It is also conceivable that enhanced public attention can trigger public appreciation of HCW. Furthermore, HCW could move to the political fore promoting improvements in the working conditions. Such rewarding aspects should be investigated in future studies.

#### 4.5.4 Implications for the general public

The importance of social support for mental health has been highlighted by several studies (Ko et al., 2006; Lau et al., 2006; Cao et al., 2020). Digital communication with friends, relatives and colleagues might buffer the negative effects of loneliness and separation. Although most of the studies have highlighted stressors and protective factors to cope with these stressors, there might even be rewarding aspects in times of an epidemic. Some positive mental health-related factors like family support, mental health awareness and lifestyle changes such as time to rest, to relax or to exercise have already been investigated (Lau et al., 2006). During epidemics, a substantial proportion of individuals might be confronted with altered working conditions like teleworking, which is generally associated with pros and cons for mental health (Mann and Holdsworth, 2003). Future studies should examine ways to reduce the negative impact of home-office situations in times of an epidemic crisis.

#### 4.5.5 Information policies for public crisis management

Many studies have highlighted the role of timely and adequate information that should be provided (Wang et al., 2020a). Epidemics with escalating case numbers and mass quarantine convey the impression of a serious personal threat and increase feelings of anxiety, loss of control and being trapped (Rubin and Wessely, 2020). The extensive mass and social media coverage is associated with public concerns and may contribute to negative psychological effects (Rubin et al., 2010; Bo et al., 2020). Appropriate information and education programs may not only help to decrease anxiety (Chan et al., 2007) but also benefit in adopting protective measures (Leung et al., 2009). Thus, adequate media is essential for the promotion of protective measures (Rubin et al., 2010). Besides the responsibility of (health-) authorities to provide adequate information, it is necessary to understand the development of public attitudes to better target communication strategies, particularly with the rise of fake news and conspiracy theories (Atlani-Duault et al., 2020). Furthermore, strengthening health literacy (Kickbusch, 2001) appears to be important in enabling people to evaluate the relevant information. Generally, the application of health behavior theories in research of public attitudes and behaviors would enhance the development of public health interventions that address the mental health-impairing processes of an epidemic crisis.

#### 4.5.6 Addressing the needs of subpopulations in public health policy

With regard to the general public, the consideration of subpopulations was mainly neglected. For instance, people with mental illness (Holmes et al., 2020) or children and families that might be victims of domestic violence, particularly in times of quarantine (Campbell, 2020). Also, for the elderly, the effects of social distancing could lead to isolation, loneliness and severe mental health consequences (Newman and Zainal, 2020). It is generally accepted to assume that people lacking resources (such as financial, cultural or social resources) might be more vulnerable within a crisis (Hobfoll, 2001). Given this, future studies should examine mental-health effects for specific subpopulations. This would result in targeted interventions in these populations in addition to general public mental health approaches.

### 4.6 Strengths and Limitations

An important strength of our study is the inclusion of a broad range of populations that may be affected by MHP during or after an epidemic. This review provides an essential overview of a highly relevant public health topic since the impact of impaired mental health itself on individuals, society and economy can be substantial. Furthermore, the data shown (Tables 1–3) allows for further interpretations and delivers insights to aspects that are of interest for researchers, practitioners and policy planning (e.g. country specific prevalence rates). Limitations may arise from our search strategy since we searched for scientific publication on PubMed and did not screen reference lists of relevant articles. Additionally, no quality assessment of the studies was conducted. Further limitations arise from the large heterogeneity and methodological issues (see sections of mental health problems and methodological issues in this paper). At the same time, the heterogeneity of integrated studies is an asset, as they offer an extensive perspective on the studied issue.

### 5 Conclusion

In this rapid review of 74 original articles, we found a large range in prevalence rates of MHP such as anxiety, depression, post-traumatic stress symptoms or disorders, during and after epidemics across the general public, HCW, and survivors. MHP might be especially prominent among HCW and survivors that are directly affected by epidemics and face a real threat of infection and difficult circumstances like isolation/quarantine or difficult working conditions. As shown by various original studies, MHP across all populations can be substantially influenced by risk and protective factors, some of which are modifiable like social support and appropriate information by authorities. From a clinical point of view, policy makers and health care providers should be aware of potential short term or even persistent MHP. During epidemics, mental health care needs to be adapted to changing circumstances in order to grant access and treatment to those in need. Digital mental health approaches can support access to care for the public. This allows for psychological monitoring and treatment when in-person consultations are not possible. Yet, digital health interventions are still in developmental stages and need further assessment. During lockdowns, they seem to be a relevant supplement to the provision of inperson mental health care. Furthermore, HCW that often account for a substantial fraction of virus cases need to be supported. However, health authorities and policy makers should keep in mind separating short-term acute stress reactions from long-term mental illness.

It is of note that many original studies used different approaches and show methodological diversity in the assessment of MHP, which at least partly explains the broad range of MHP. Thus, results should be treated with some caution since a comparison of prevalence rates across studies and assessment of magnitude of MHP is currently not possible. Future studies should monitor MHP with standardized methods and apply comparisons with country-specific norms in order to gain a better understanding of MHP, to learn about influential factors, and to better understand how to provide appropriate access to mental health care during epidemics. Although, this was out of scope for this review, evidence of MHP in vulnerable populations such as children or people with pre-existing mental illness seems to be scarce and should be covered in future studies.

## Data Availability

not applicable

## 7 Author Contributions

DR and SJZ contributed to the design of the study, data acquisition, data interpretation, manuscript development and revisions. PK contributed to data acquisition, data interpretation, manuscript development and revisions. CA, FH contributed to data acquisition and manuscript revisions. CB contributed to data interpretation and manuscript revision. AL contributed to data interpretation, manuscript development and revisions. All authors approved the final version of the submitted manuscript.

## 9 Acknowledgements

No acknowledgements

## 10 Supplements

**Supplementary Table 1.** PubMed search strategy (search at April 1, 2020)

|  |  |
| --- | --- |
| Population | Sars*[Title/Abstract] OR COVID*[Title/Abstract] OR 2019-nCoV[Title/Abstract] OR Corona[Title/Abstract] OR Coronavirus[Title/Abstract] OR Mers*[Title/Abstract] OR “Severe Acute Respiratory”[Title/Abstract] OR Influenza*[Title/Abstract] OR Flu[Title/Abstract] OR Grippe[Title/Abstract] OR Ebola*[Title/Abstract] |
| AND |  |
| Outcomes | Mental[Title/Abstract] OR Psychological[Title/Abstract] OR depress*[Title/Abstract] OR schizophren* OR psychosis OR psychotic OR anxiety OR bipolar OR Stress[Title/Abstract] OR PTSD [Title/Abstract] OR Emotional [Title/Abstract] OR Trauma*[Title/Abstract] |
| NOT |  |
| Conditions / Interventions | Coronary[Title] OR Cardiac[Title] OR Cardiovascular[Title] OR “Physical activity”[Title] OR Biomarkers[Title] OR Injury[Title] OR Anaesthesia[Title] OR Melatonin[Title] OR Validation[Title] OR Pain[Title] OR Obesity[Title] OR Opioid[Title] OR fentanyl[Title] OR Orthopaedic[Title] OR Vaccin*[Title]) |

